# Cov2clusters: genomic clustering of SARS-CoV-2 sequences

**DOI:** 10.1101/2022.03.10.22272213

**Authors:** Benjamin Sobkowiak, Kimia Kamelian, James E. A. Zlosnik, John Tyson, Anders Gonçalves da Silva, Linda M. N. Hoang, Natalie Prystajecky, Caroline Colijn

## Abstract

**Background:** The COVID-19 pandemic remains a global public health concern. Advances in sequencing technologies has allowed for high numbers of SARS-CoV-2 whole genome sequence (WGS) data and rapid sharing of sequences through global repositories to enable almost real-time genomic analysis of the pathogen. WGS data has been used previously to group genetically similar viral pathogens to reveal evidence of transmission, including methods that identify distinct clusters on a phylogenetic tree. Identifying clusters of linked cases can aid in the regional surveillance and management of the disease. In this study, we present a novel method for producing stable genomic clusters of SARS-CoV-2 cases, cov2clusters, and compare the sensitivity and stability of our approach to previous methods used for phylogenetic clustering using real-world SARS-CoV-2 sequence data obtained from British Columbia, Canada,

**Results:** We found that cov2clusters produced more stable clusters than previously used phylogenetic clustering methods when adding sequence data through time, mimicking an increase in sequence data through the pandemic. Our method also showed high sensitivity when compared to epidemiologically informed clusters.

**Conclusions:** Our new approach allows for the identification of stable clusters of SARS-CoV-2 from WGS data. Producing high-resolution SARS-CoV-2 clusters from sequence data alone can a challenge and, where possible, both genomic and epidemiological data should be used in combination.

## Background

The COVID-19 pandemic has had worldwide economic, social and health impacts unlike any infectious disease in recent history. First identified as an unknown cause of pneumonia in patients from Wuhan, China in late 2019, the aetiological agent was quickly determined to be a novel *Betacoronavirus*, subsequently named severe acute respiratory syndrome coronavirus 2 (SARS-CoV-2) ^1–3^. Extensive global person-to-person transmission followed and on March 11, 2020 ^4^, the World Health Organization (WHO) declared COVID-19 a pandemic, with cases since reported in almost every country in the world. As of 10^th^ March 2022, there have been over 450 million cases and 6 million deaths associated with the disease worldwide ^5^.

The development of effective vaccines and regional containment strategies have allowed countries to mitigate the spread of SARS-CoV-2 and thereby reduce transmission, hospitalization, and death rates from COVID-19. Nevertheless, the threat posed by the disease is still a worldwide concern due to the emergence of Variants of Concern (VoCs) such as the Delta and Omicron variants that display increased transmissibility with lower vaccine effectiveness ^6,7^, delayed global vaccination deployment, vaccine hesitancy, and unequal access to vaccines and therapeutics.

We have seen an unparalleled effort in whole genome sequencing (WGS) of COVID-19 to identify new variants and mutations of concern. To date, there are over 9 million sequences publicly available through the open-source GISAID initiative ^8^. Utilising these data to develop novel and easy-to-implement tools to detect growing or emerging transmission clusters can help control the spread of the virus locally. We can use genomic similarity to identify linked cases with shared demography or geography at a higher resolution than a shared lineage assignment or simply via contact tracing. Inspecting clusters can reveal sources of common exposures or patterns of transmission through a population, which can be used to understand regional epidemiology and inform public health policy, such as implementing restrictions in certain settings with a high transmission risk. Practically, we have also seen that the SARS-CoV-2 lineage nomenclature, such as the widely used Pangolin system ^9^ has been dynamic through the pandemic and cannot provide sufficient resolution for epidemiological investigations. Thus, clustering sequences by genomic similarity provides the resolution and stability necessary for public health applications over the course of a dynamic pandemic.

Phylogenetic trees are an effective tool for summarizing evolutionary relationships among taxa, and tree reconstruction methods can be used to achieve realistic measures of genetic divergence. The information contained within a phylogeny can be used to define groups of closely related sequences that may indicate recent transmission between cases, either through identifying distinct clades on a tree or by using the pairwise patristic distance as a measure of divergence between tips. Phylogenetic clustering methods have been applied in many virological analyses ^10–12^, as well as early in the COVID-19 pandemic to define putative transmission clusters in SARS-CoV-2 ^13–15^. However, clustering based solely on genetic variation may not be sufficient to effectively identify meaningful clusters in SARS-CoV-2 where there has been rapid spread of the virus with relatively low genetic diversity ^16–18^, as well as periods of lineage replacement with new VoCs also reducing regional genetic diversity in the virus ^19^. Additionally, comprehensive sampling of ongoing transmission within a population can result in multiple clusters that are linked genetically through ancestral samples. Defining clusters using a fixed genetic distance threshold may cause sequences to change cluster designation through time as more sequences are collected.

Here, we present a novel method for constructing SARS-CoV-2 genomic clusters, using the pairwise probability of clustering under a logit regression model, and linking cases under a given probability threshold. The logit model incorporates genetic relatedness through phylogenetic distance and collection or symptom onset date; this method also allows for the inclusion of other covariates of interest that may result in meaningful clusters (e.g., contact data, exposure events). In contrast to previous clustering approaches that often rely solely on phylogenetic inference (tree cluster reference), clustering isolates in this pairwise manner allows for greater cluster stability through time, as well as resolution by including epidemiological information without the need for time-consuming manual investigation. Previous clustering designation of sequences can also be specified *a priori* to further improve cluster stability. This also allows clustering to be performed on subsampled datasets where previously clustered sequences have been removed for ease of analysis. We provide this method as an R package, github.com/bensobkowiak/cov2clusters, for use within the research and public health community to investigate SARS-CoV-2 transmission dynamics.

## Results

### Sample description

Whole genome sequence data was obtained for 36,420 SARS-CoV-2 samples collected between 15^th^ March 2020 and 13^th^ August 2021 in BC, Canada. These data encompass sequences collected during the first, second and third ‘waves’ of the pandemic in the province, predominantly comprising the SARS-CoV-2 sub-lineages B.1.2 and B.1.438.1 (wave 2) initially before replacement with B.1.1.7 (Alpha) and P.1 (Gamma) (wave 3), which led to a rapid increase in cases (**Figure 1A)**. The data also includes the beginning of a rise in cases in June 2021 that would eventually lead to the fourth ‘wave’ in BC, largely driven by the Delta SARS-CoV-2 variant sub-lineages B.1.617.2 and AY.25 (more recently denoted as AY.25.1), with the number of cases caused by this variant increasing in early May 2021 before becoming the principal variants in BC by August 2021 (**Figure 1B**). Levels of genetic diversity within the SARS-CoV-2 samples collected fluctuated over the study period, with very low diversity in the population observed during the periods of high B.1.1.7 (Alpha) and P.1 (Gamma) case numbers, and with the introduction of the Delta variants (**Supplementary figure S1)**.

**Figure 1.**
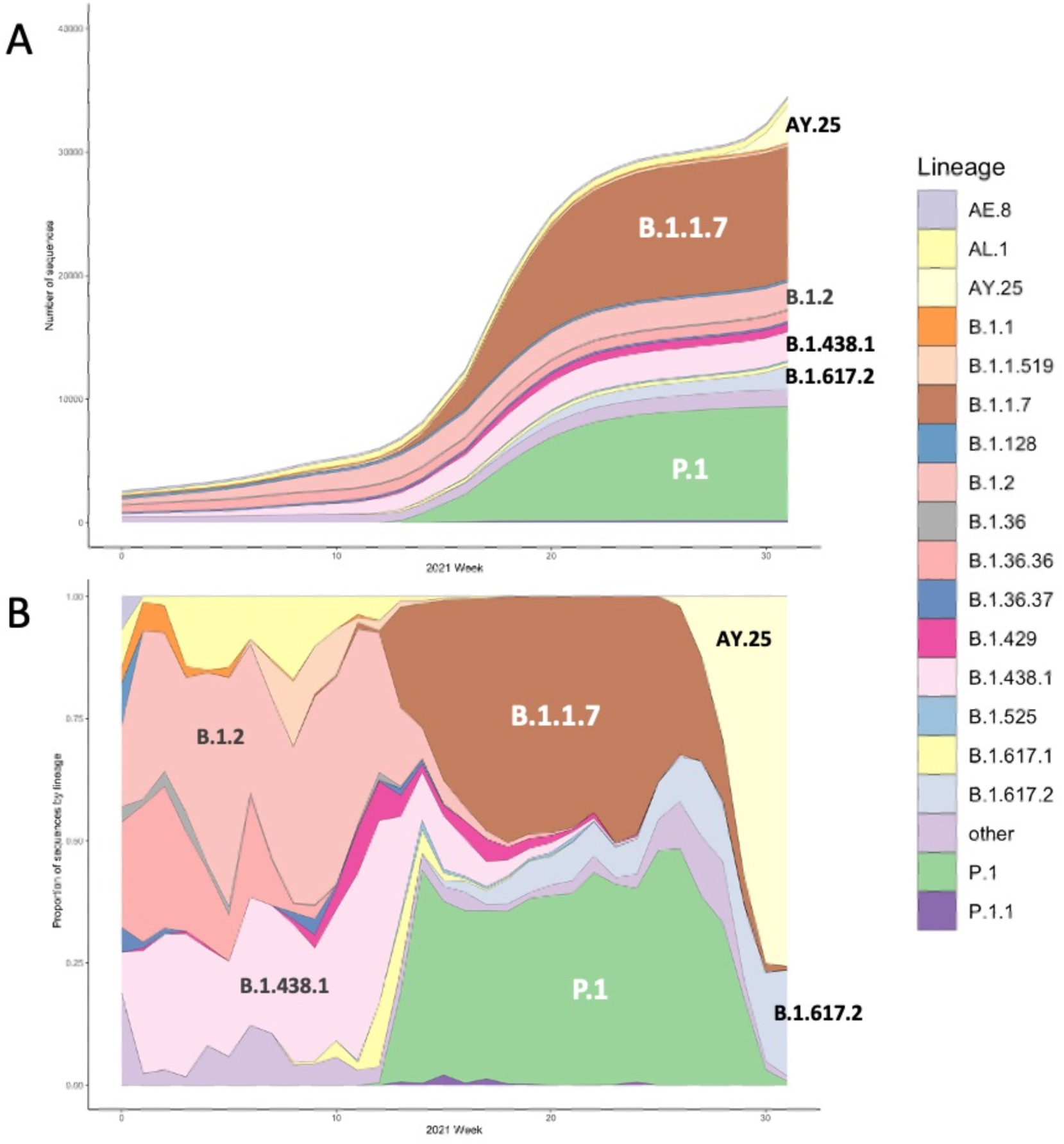
The cumulative number (A) and lineage proportion (B) of SARS-CoV-2 sequences per week included in the study, coloured by lineage. Major lineages present in the data are annotated.

### Cluster results in BC SARS-CoV-2 data

Genomic clustering with cov2clusters (using the pairwise probability thresholds of 0.7 and 0.8) and TreeCluster ‘single_linkage’ found fewer, larger clusters than both cov2clusters at the 0.9 threshold and TreeCluster ‘max_clade’. This occurs both in the pre-Delta dominance and Delta wave data. cov2clusters at the 0.9 probability threshold found many small clusters and a high number of sequences assigned as non-clustered, indicating this threshold may over-cluster the data. **Figure 2** shows the phylogenetic trees produced by the pre-Delta dominance and Delta waves and the resulting cluster assignments, with the largest five clusters found by each approach shown in colours (cluster size range N = 194 - 4638 pre-Delta dominance, and cluster size range N = 181 – 4323 Delta wave), all sequences clustered in smaller clusters in grey, and non-clustered sequences in white. The largest clusters found using cov2clusters at the 0.7 and 0.8 probability thresholds were of similar size (N = 4452 – 4638 pre-Delta dominance, and N = 4250 - 4323 Delta wave) though the number of clusters at the 0.7 probability threshold was lower (567 vs 756 clusters pre-Delta dominance, and 453 vs 591 clusters Delta wave), with most sequences of the same sub-lineage assigned to a single, large cluster.

**Figure 2.**
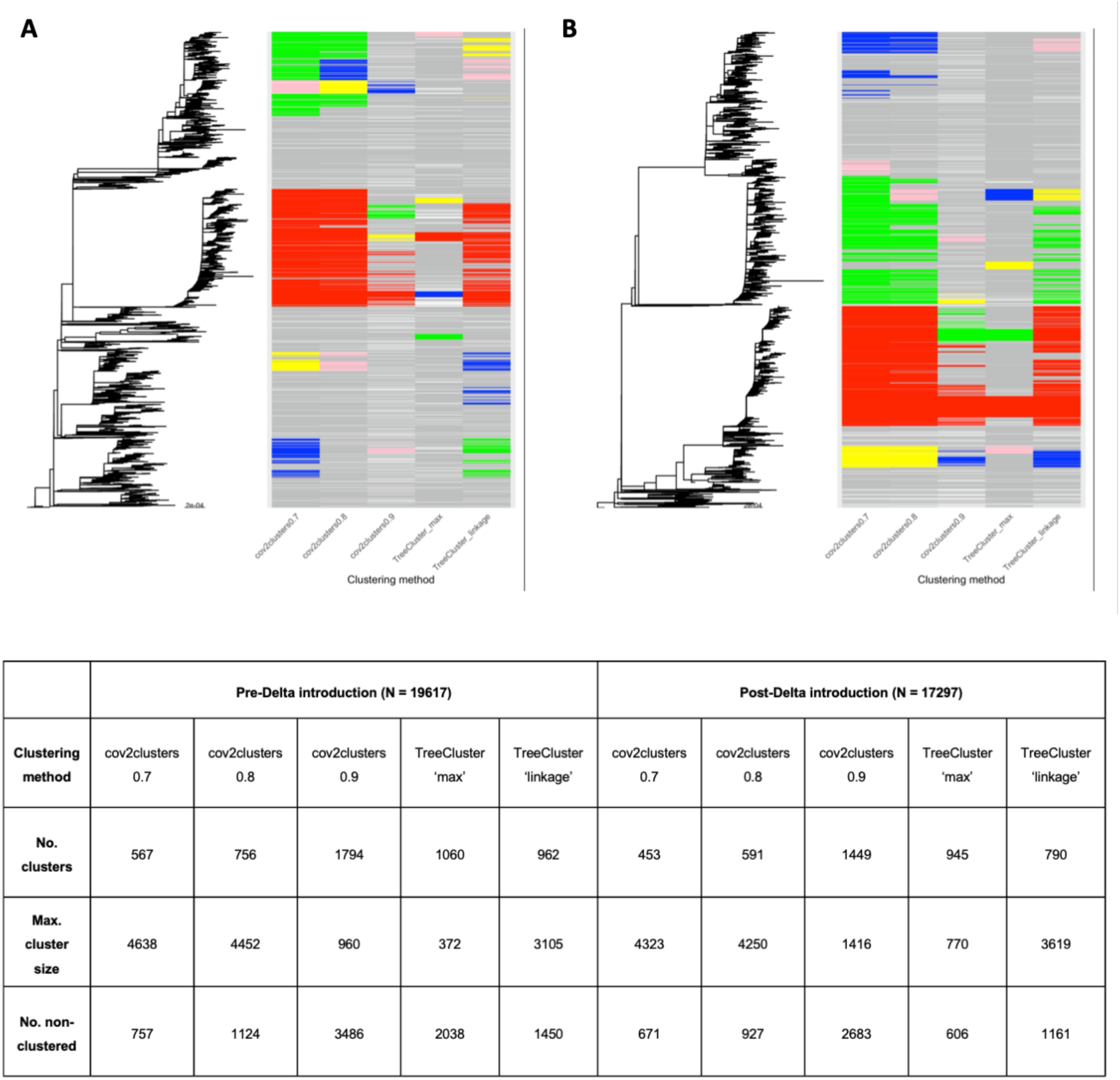
Maximum-likelihood phylogenies and clustering assignments of the (A) pre-Delta dominance wave (N = 19,617) and (B) Delta wave (N = 17,297) sequences. Sequences in the largest five clusters found by each method are coloured, with those in the largest cluster in red, followed by green, blue, yellow and pink. All other clustered sequences are coloured grey, and non-clustered sequences are in white.

### Sensitivity of clustering methods with epidemiologically informed clusters

The sensitivity of the logit clustering tool, cov2clusters, for assigning sequences into seven epidemiologically well-defined clusters was tested using two pairwise probability thresholds (0.8, 0.9). These results were also compared to the sensitivity of TreeCluster using both the maximum clade distance threshold approach and the maximum pairwise linkage threshold approach (**Table 1**). We found that cov2clusters with a pairwise probability threshold of 0.8 assigned the highest number of sequences to epidemiologically informed clusters (92%), performing marginally better than TreeCluster ‘single_linkage’ method (87%) and significantly better than TreeCluster ‘max_clade’ (66%) and cov2clusters at the 0.9 probability threshold (71%). We also tested cov2clusters with a probability threshold of 0.7, though at this threshold very large lineage-specific clusters were formed, which did not add any meaningful resolution to transmission clusters compared to simply using lineage classification.

**Table 1.**
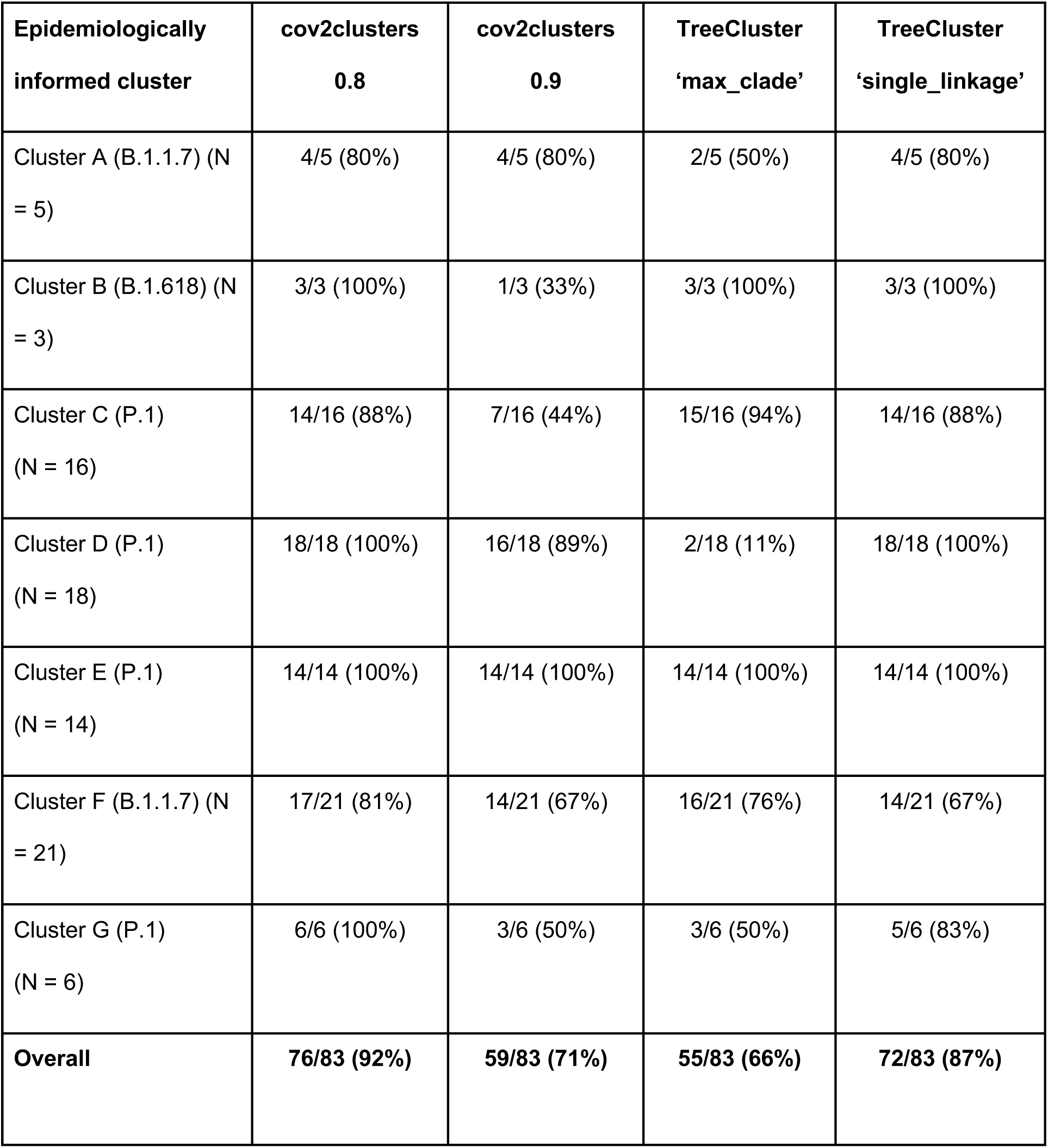
The sensitivity of cov2clusters at three pairwise probability thresholds (0.8 and 0.9) and TreeCluster (‘max_clade’ and ‘single_linkage’ methods) for placing sequences within seven epidemiologically well supported SARS-CoV-2 clusters in British Columbia, Canada.

### Cluster stability through time

The stability of the genomic clusters through time was assessed by running each method on the Delta wave data collected up to 11^th^ June 2021, and then re-running the clustering, adding sequences collected each subsequent week until the end of the study period. Stability measures tested were 1) the proportion of sequences that moved from a cluster in the preceding week to non-clustered in the current week, 2) the number of clusters defined in the previous week that split in the current week (i.e., any instance where sequences that were in a single cluster in the previous week have moved to different clusters in the current week), and 3) the overall entropy score of the clusters found in the current week (with the lowest score of 0 occurring when all sequences are in a single cluster).

We found that the TreeCluster ‘max_clade’ method resulted in the highest proportion of sequences moving from clusters to become non-clustered in subsequent weeks (highest on 23^rd^ July 2021 with 1.14% of sequences). TreeCluster ‘single_linkage’ resulted in lower numbers of sequences moving from clustered to non-clustered (**Figure 3)**. All cov2clusters methods did not result in any sequences moving from clustered to non-clustered. The number of cluster splits was also highest with TreeCluster ‘max_clade’, with 54 clusters splitting in the week ending 13^th^ August 2021, followed by TreeCluster ‘single_linkage’ with 9 clusters splitting in the week ending 23^rd^ July 2021. Cluster entropy was observed at its lowest in cov2clusters at the 0.7 threshold through every week of the tested period, followed by cov2clusters at the 0.8 threshold. cov2clusters at a 0.9 threshold and TreeCluster ‘max_clade’ scored the highest entropy, reflecting the more even distribution of the data into smaller clusters, though as shown, this can be at the cost of over-clustering.

**Figure 3.**
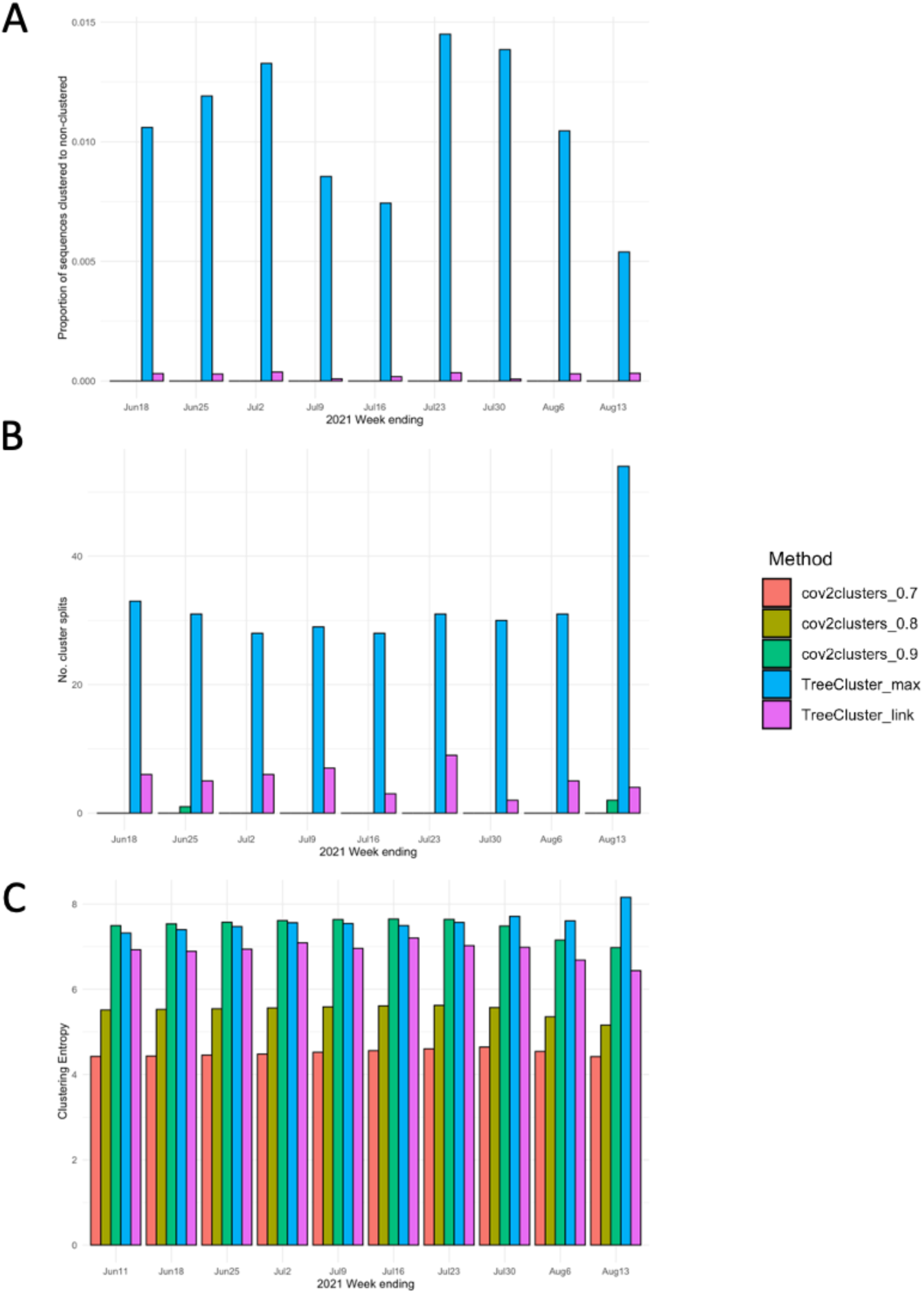
Cluster stability for each tested method assessed on SARS-CoV-2 sequences collected in British Columbia, Canada before 18^th^ June 2021, adding sequences collected each week until 13^th^ August 2021. (A) The proportion of sequences becoming non-clustered when clustered the week previously, (B) the number of cluster splits, and (C) the clustering entropy score.

## Discussion

In this study, we have presented a new method for genomic clustering of SARS-CoV-2 using pairwise probabilities of shared cluster membership derived from a logit regression model based on sequence divergence and sample collection dates. This method can readily incorporate epidemiological data, such as geography, contact or shared exposure, to add further resolution to clustering. We tested our approach using three pairwise probability thresholds for linking sequences to form clusters and found that at probability threshold of 0.8 formed the most stable clusters in our clinical data from samples collected in BC, Canada. Comparing our method to other phylogenetic clustering tools, we found the sensitivity of cov2clusters at a 0.8 probability threshold was higher than both TreeCluster ‘max_clade’ and ‘single_linkage’, and that the clusters produced were more stable as cases are added through time, owing to our approach incorporating past designations in time into the clustering pipeline. This result has particular significance for the utility of this method in real-time public health surveillance, where sequencing datasets grow over time, and stability in cluster designations is beneficial for reporting and surveillance. We have implemented our approach as a freely available R package.

We used patristic distance from phylogenetic trees as the measure for genetic divergence in our method to utilize the full information available in the sequence alignment, compared to genetic distance measures that, while correlated with patristic distance, may underestimate pairwise divergence ^20^. Phylogenetic uncertainty in SARS-CoV-2 trees, where many terminal branches are supported by low numbers of mutations, has been explored previously ^21^. It was shown that variation in tree topology, which in turn will alter pairwise distances between tips, was driven by the sample set of sequences used to construct the tree that changes through time. While this will impact the stability of any method that uses patristic distance to inform clustering, we have shown that our approach reduces this instability in genomic clustering.

Large clusters of genetically similar sequences were common in our dataset. Indeed, given the high number of COVID-19 infections and relatively low genetic diversity of SARS-CoV-2 in the province, it is expected that in settings with even moderate levels of sequencing, we are likely to capture sequences separated by few mutations. Therefore, large clusters will occur, with many identical or near-identical sequences and with ‘chaining’ of closely related sequences. In other words, with dense sampling of ongoing person-to-person transmission, and over a short timeframe, there may be a lack of well-separated clusters in datasets for any clustering method to uncover. This contrasts with some other viruses, such as HIV, that will produce structured phylogenies from which discrete clusters can be identified ^20^. This in part due to HIV’s chronic nature (leaving longer time intervals between infections with a higher potential for intra-host genetic diversity and viral populations), as well as the fact that, in HIV, relatively small clusters are seeded by introductions from other jurisdictions. Here, when a large fraction of infections is sequenced, the time between infections is short, and considerable transmission is occurring within the sampling jurisdiction. Therefore, using only genomic divergence derived from a given phylogeny is unlikely to identify well-separated SARS-CoV-2 transmission clusters. Additional epidemiological data can be used to refine large clusters found using our genomic clustering approach. For example, including information such as common exposures and contact tracing data may divide large clusters into operational units with public health relevance. One limitation of our study is that we do not have exposure, contact or location information to explore this application.

Sequences belonging to a P.1 sublineage (P.1.14) form a single, large cluster (illustrated as the red cluster in the delta wave dataset in **Figure 2**), coinciding with a high number of low-diversity P.1 cases present in BC from April 2021 onwards ^22^, where almost all P.1 samples were within 0-1 SNPs of another P.1 sequence. This phenomenon is also expected with the recent Omicron variant, where rapid spread has led to high numbers of low diversity cases ^23^. Increasing the probability threshold to 0.9 (or conducting phylogenetic clustering with a smaller maximum clade divergence threshold) breaks up the cluster into smaller groups of identical or near-identical sequences, but this does not reflect genuine underlying clustering (**Supplementary figure S2**). In such circumstances, we recommend including additional metadata to refine clusters into genetically related groups with shared demography and epidemiology. Alternatively, our approach could be used as a surveillance tool focusing on a particular individuals or settings of interest, identifying sequences that are linked to the focal individuals or exposure sites, moving outwards to a desired number of “rings”.

While COVID-19 remains at pandemic levels with high case numbers in many regions globally, it is anticipated that there will be a shift to endemicity characterized by persistent, lower levels of the disease interspersed with seasonal or occasional outbreaks ^24^. In that context, it is likely that the viral population will have smaller and better-separated clusters. We suggest that the method presented here for clustering can be effectively utilized in both contexts.

## Conclusions

Identifying meaningful, high-resolution clusters from SARS-CoV-2 genomic sequence data alone can be a challenge due to relatively low genetic diversity and high rates of localised transmission. Nevertheless, WGS data can be a useful tool when combined with epidemiological and demographic data to better characterise groups of individuals with shared transmission histories. Here we present a simple method for producing highly stable genomic clusters of SARS-CoV-2 that incorporates genomic and epidemiological data to link cases for use in public health surveillance.

## Methods

### Sequence data and phylogenetic analysis

Positive SARS-CoV-2 samples collected in British Columbia (BC), Canada, between 18^th^ March 2020 and 13^th^ August 2021 underwent whole-genome sequencing at the BCCDC Public Health Laboratory. Sequencing sampling strategy changed over the course of the pandemic and increases with sequencing capacity at the lab. Sampling strategies included random sampling (ranging from 5-100% of cases at different periods) and targeted sampling (outbreaks and targeted populations such as travellers)^25^. Sequence data used in this study have been deposited in the GISAID database ^8^.

Nucleic acids were extracted using the MagMAX instrument from Thermofisher (AM1836) and amplified using the Freed primer scheme (1200 base pair amplicons) detailed here ^26^. Consensus sequences were generated using the Connor Laboratory pipeline (https://github.com/connor-lab/ncov2019-artic-nf) with consensus bases called at a frequency of 0.75 with a subsampling read count strategy. Consensus sequences were aligned and trimmed to Wuhan-Hu-1 reference sequence (Accession MN908947, Version MN908947.3) using MAFFT (v7.471) ^27^ prior to phylogenetic tree production. A specific fork of the ARTIC pipeline for processing SARS-CoV-2 sequences was created to support the SARS-CoV-2 sequencing efforts at the BCCDC Public Health Laboratory, located here: https://github.com/BCCDC-PHL/ncov2019-artic-nf. Sequences with no collection date or excess or ambiguous sites (> 15% missing calls) were removed from the analysis.

### Phylogenetic analyses

A multiple sequence alignment of the full SARS-CoV-2 genome was used to construct maximum-likelihood (M-L) phylogenetic trees with IQ-TREE (v.2.1.3) ^28^. One sequence per individual was included for analysis, with the earliest sequence chosen where longitudinal samples were taken from the same disease episode. Optimal nucleotide substitution models for the data were chosen using ModelFinder in IQ-TREE ^29^ and applied to each tree construction pipeline. For comparison to the proposed clustering approach, phylogenetic clustering was performed using TreeCluster (v.1.0.3) ^30^ using two thresholds, 1) a maximum divergence threshold within clusters of 4×10^−4^ substitutions/genome (used previously for generating SARS-CoV-2 phylogenetic clusters ^13^), and 2) a maximum pairwise divergence threshold (among pairs in a cluster) of 5×10^−5^ substitutions/genome, equivalent to a SNP distance of between 1-2 SNPs.

### Genomic clustering methodology

Genomic clusters were defined as networks of connected sequences (nodes) where the pairwise probability of clustering was above a given threshold. The probability of clustering between two sequences was calculated using the logit model:

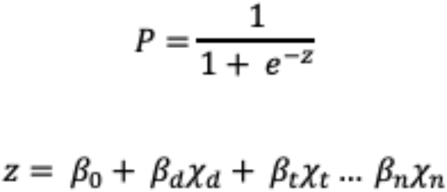

Coefficients (*β*) can be either manually chosen or estimated using the logistic regression on data with known clusters. *d* is the pairwise genetic divergence, calculated between all pairs of isolates by extracting patristic distances (the sum of branch lengths connecting two tips) on the phylogenetic tree of up to 40,000 sequences (the upper computational limit), in units of substitutions/genome. *t* is a measure of difference in time between sequences, either the date of collection or symptom onset, and can be extracted from the associated metadata or inferred from a timed phylogeny. Additional covariates (*n*), such as contact data between hosts or shared exposure events, can be included to further resolve clusters. Pairwise transmission probabilities calculated in previous clustering runs can be included in new analysis to allow for greater continuity in cluster designations, as well as permitting subsequent clustering runs to be run on subsampled datasets to increase speed and efficiency when clustering large numbers of sequences. The full R code is available in the supplementary materials and available at github.com/bensobkowiak/cov2clusters.

We compared the results of our genomic clustering method at three pairwise probability thresholds of 0.7, 0.8 and 0.9 to link sequences to the clusters obtained using TreeCluster ‘max_clade’ (where the maximum pairwise patristic distance threshold between any two sequences in a cluster was 4 × 10^4^ substitutions per sequence site) and ‘single_linkage’ (where any two sequences up to a maximum patristic distance threshold of 5 × 10^5^ substitutions per sequence site must be in the same cluster). The data were separated into two large datasets, defined as pre-Delta dominance wave (N = 19,617), which included all sequences collected before 6^th^ May 2021, and Delta wave (N = 17,297), which included all sequences after this date, as well as 500 randomly selected sequences collected before this date as a representative skeleton tree of past diversity. Beta coefficients fo r the genomic clustering algorithm of *β*_*0*_ = 3, *β*_*1*_ *=* -1.9736 × 10^−4^, and *β*_*2*_ = 7.5 × 10^−2^ were manually chosen, which corresponds to a pairwise probability of 0.95 between sequences with the same genomic sequence and collected date, with a decrease in pairwise probability as the patristic distance and/or collection date difference increases. **Supplementary figure S3** provides the results of the pairwise probabilities using logistic regression with these beta coefficients with variable patristic distance (converted to SNP distance by multiplying by the genome length) and difference in collection dates between sequences.

## Data Availability

Whole genome sequence data included in this study are deposited in the GISAID repository https://www.gisaid.org. The cov2clusters code is available at: https://github.com/bensobkowiak/cov2clusters.

## Ethics approval and consent to participate

The study was approved by the University of British Columbia Ethics Board (#H20-02285).

## Consent for publication

Not applicable.

## Competing interests

The authors declare they have no competing interests.

## Funding

This work was supported by funding from Michael Smith Foundation for Health Research and he Federal Government of Canada’s Canada 150 Research Chair program. This work was supported by the Canadian Covid genomics network (CanCoGen).

## Authors’ contributions

BS, CC, and NP conceived the project. BS and CC developed the methodology, with support from KK, JEAZ, AGdS and NP. Data were provided by KK, JT, LMNH and NP. BS performed the analysis. BS and CC wrote the first version of the manuscript, and KK, JEAZ, LMNH, and NP provided revisions for the final manuscript. All authors have read and approve of the final version of the manuscript.

## Acknowledgements

We would like to acknowledge the work of the Public Health Laboratory at the British Columbia Centre for Disease Control (BCCDC), for providing the SARS-CoV-2 isolates used in this study

## Supplementary Figures

**Figure S1.**
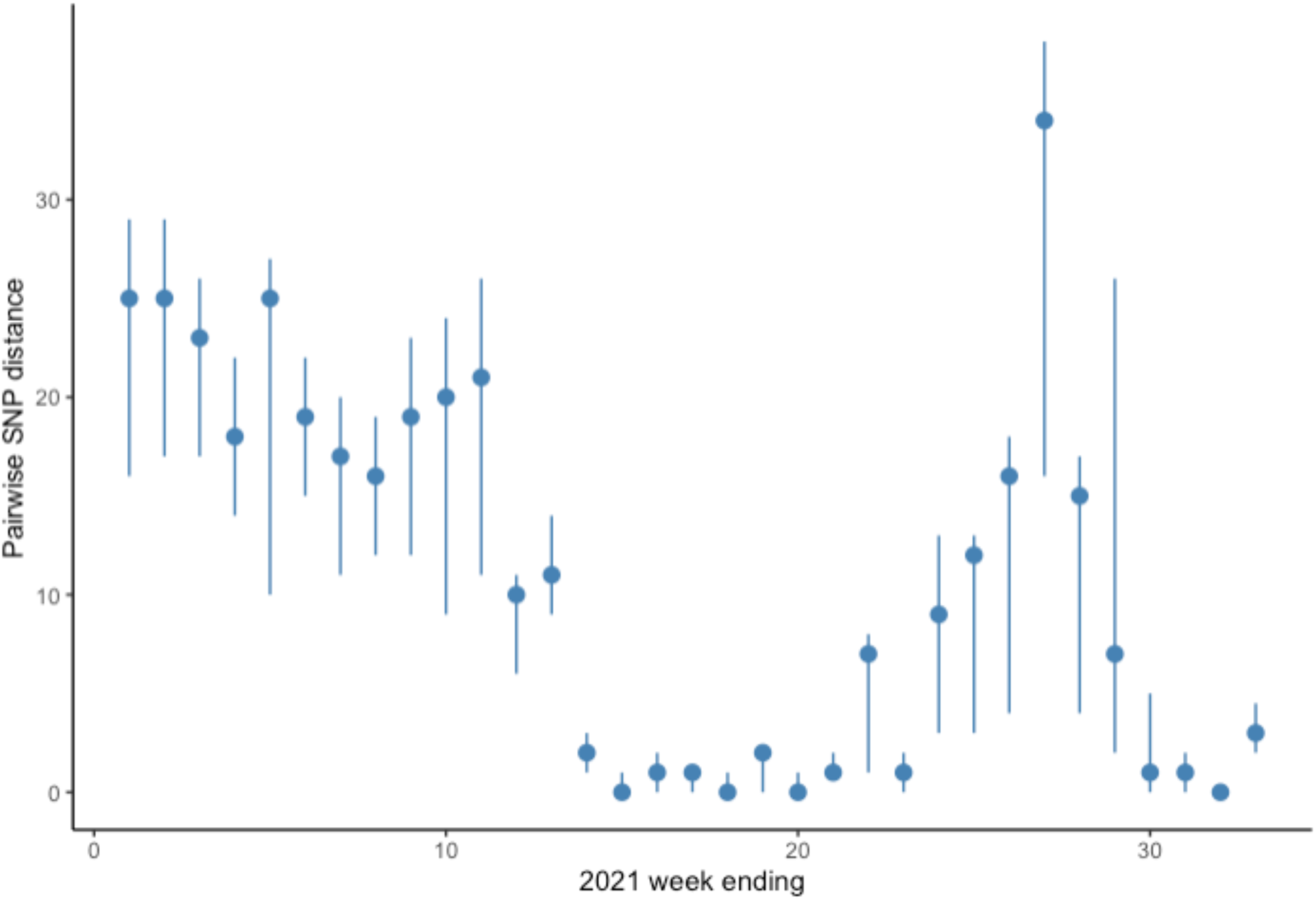
Median and interquartile pairwise SNP distance between sequences collected in the study period by week of 2021.

**Figure S2.**
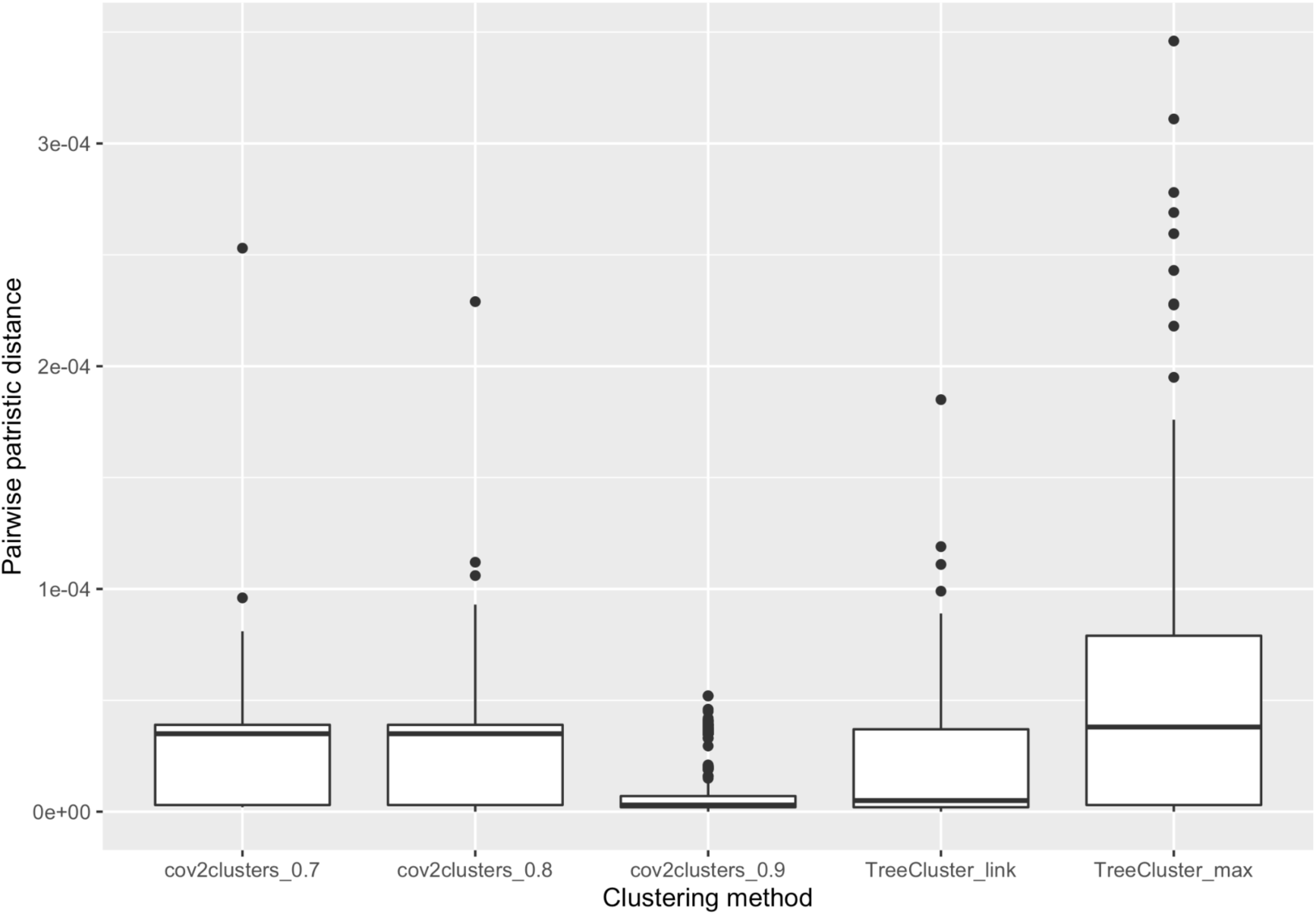
The pairwise patristic distance between P.1 sequences in clusters identified by each clustering approach.

**Figure S3.**
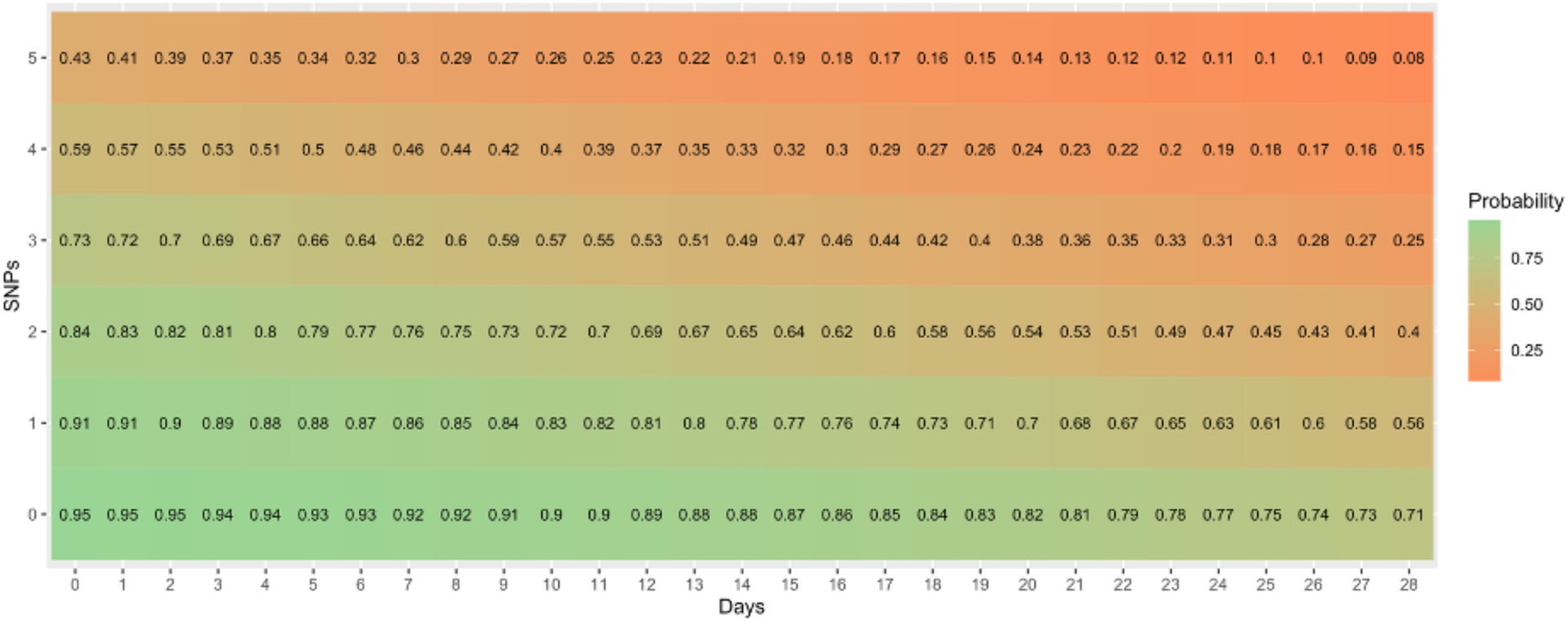
The pairwise probability of linking two sequences by SNP distance and difference in collection date using the selected beta coefficients used in this study (= 3, = -1.9736 × 10^−4^, and = 7.5 × 10^−2^). Patristic distance has been converted to SNP distance by multiplying SNP distance by the genome length for easier interpretation of pairwise sequence distance.

## Notes

### Competing Interest Statement

The authors have declared no competing interest.

### Author Declarations

The study was approved by the University of British Columbia Ethics Board (#H20-02285).

### Summary of Updates

Results updated with figures quoted

